# Defining the Pandemic at the State Level: Sequence-Based Epidemiology of the SARS-CoV-2 virus by the Arizona COVID-19 Genomics Union (ACGU)

**DOI:** 10.1101/2020.05.08.20095935

**Authors:** Jason T. Ladner, Brendan B. Larsen, Jolene R. Bowers, Crystal M. Hepp, Evan Bolyen, Megan Folkerts, Krystal Sheridan, Ashlyn Pfeiffer, Hayley Yaglom, Darrin Lemmer, Jason W. Sahl, Emily A. Kaelin, Rabia Maqsood, Nicholas A. Bokulich, Grace Quirk, Thomas D. Watts, Kenneth Komatsu, Victor Waddell, Efrem S. Lim, J. Gregory Caporaso, David M. Engelthaler, Michael Worobey, Paul Keim

## Abstract

In December of 2019, a novel coronavirus, SARS-CoV-2, emerged in the city of Wuhan, China causing severe morbidity and mortality. Since then, the virus has swept across the globe causing millions of confirmed infections and hundreds of thousands of deaths. To better understand the nature of the pandemic and the introduction and spread of the virus in Arizona, we sequenced viral genomes from clinical samples tested at the TGen North Clinical Laboratory, provided to us by the Arizona Department of Health Services, and at Arizona State University and the University of Arizona, collected as part of community surveillance projects. Phylogenetic analysis of 79 genomes we generated from across Arizona revealed a minimum of 9 distinct introductions throughout February and March. We show that >80% of our sequences descend from clades that were initially circulating widely in Europe but have since dominated the outbreak in the United States. In addition, we show that the first reported case of community transmission in Arizona descended from the Washington state outbreak that was discovered in late February. Notably, none of the observed transmission clusters are epidemiologically linked to the original travel-related cases in the state, suggesting successful early isolation and quarantine. Finally, we use molecular clock analyses to demonstrate a lack of identifiable, widespread cryptic transmission in Arizona prior to the middle of February 2020.

## Introduction

In late 2019, a novel positive sense RNA virus, family *Coronaviridae*, genus *Betacoronavirus* and subgenus *Sarbecovirus*, emerged in the human population as a result of cross-species transmission from an unknown host^1,2^. The virus, SARS-CoV-2, began widespread circulation in the Chinese city of Wuhan in late December of 2019 with spread in other countries apparently beginning around mid-January^3,4^.

The first confirmed case of COVID-19, the disease caused by SARS-CoV-2, in Arizona was in late January 2020 from a student attending Arizona State University who had travelled to China^5^. Extensive contact tracing and isolation led to zero additional reported cases stemming from this original case (AZ1). There were no additional cases reported in Arizona until March 3rd, when a travel-related case returned from Europe and tested positive^6^. On March 6th, the first case of community transmission in Arizona was announced^7^. On March 26th, the Arizona Department of Health Services (AZDHS) updated the level of community transmission to “Widespread.” As of May 6th, there are ~9,707 confirmed positive cases in Arizona and more than 426 deaths^8^, and the number of new cases per day in Arizona continues to increase^9^. The Navajo Nation, located mostly in northeastern Arizona (but also Utah, Colorado, and New Mexico), at the time of writing has among the highest number of cases per capita in the US: approximately 850 confirmed cases per 100,000 individuals^10^. This is disproportionately high relative to the rest of Arizona (approximately 130 confirmed cases per 100,000 individuals), and about 50% of the per capita cases seen in the hardest hit regions of the USA (areas of New York^11^).

Given the scale of the pandemic, there is an urgent need to understand patterns of SARS-CoV-2 spread, including the relative roles of local transmission versus repeated travel-associated introductions, and the accumulation and spread of mutations that could affect the function of the virus, interfere with testing, or have antigenic effects that might impact vaccine efforts. Viral genome sequencing has emerged as a key tool for addressing these questions. In order to assist with both local and global efforts to track the spread and evolution of this virus, we began intensive sequencing of viral genomes from across Arizona and deposition of these sequences in the GISAID database, which makes them accessible to the research community for downstream analyses, notably including real-time pathogen tracking through Nextstrain^12^.

To better understand the evolution of the virus within the state of Arizona we compared our sequences with publicly available SARS-CoV-2 genomes from across the world in a phylogenetic framework, and here we report our preliminary findings. Specifically, we sought to answer 3 key questions regarding the circulation of the virus in Arizona. First, although it appeared from epidemiological data that the first case of COVID-19 in Arizona did not lead to any additional cases, did cryptic transmission occur, similar to that proposed to have begun with WA1, the first diagnosed COVID-19 patient in the U.S., who arrived in Washington state from Wuhan, China on January 15th, 2020^13^? Second, how many independent introductions contributed to the outbreak in Arizona and what was the approximate timing of each event? Third, are there any unique mutations present in Arizona sequences that could have potential phenotypic effects or could interfere with diagnostic detection?

## Results/Discussion

As of April 15th, we have sequenced and assembled a total of 79 nearly complete SARS-CoV-2 genomes obtained from patients across Arizona. These genomes were sequenced from nasopharyngeal swabs that were collected over a 28 day period from March 5th to April 2nd, 2020. This represents a sequencing effort of 4.9% of all reported cases in Arizona as of April 2nd. This dataset includes at least one genome from 11 of the 15 Arizona counties (Supplementary Table 3). Aside from AZ1, 3 SARS-CoV-2 genomes from Arizona were sequenced by the CDC, which we do not include in any analyses.

### Initial Arizona case did not lead to sustained local transmission

The first case of COVID-19 in Arizona (AZ1) was documented in late January 2020, and contact tracing suggested this initial case did not result in additional symptomatic infections within the state^5^. We sought to independently verify this conclusion using Bayesian and maximum likelihood phylogenetic analyses to compare our 79 genomes to a representative subset of the SARS-CoV-2 genomes generated from around the world. The genomes used in this analysis were selected using a novel bioinformatics pipeline (see Methods), which subsampled genomes uploaded to GISAID to reduce the size of the dataset while representing temporal, spatial, and genetic diversity of the full dataset. This resulted in a set of 384 SARS-CoV-2 genome sequences, including our 79 Arizona genomes and 305 additional representatives (Supplementary Table 1). A reduced version of this dataset, including 372 genomes with complete dating information, was used in the Bayesian analysis.

Our phylogenetic analyses (Figure 1 and Supplementary Figure 1) indicated that the genome from AZ1 belonged to lineage A. Although 9/79 (11.4%) of our genomes also clustered in lineage A, these genomes belonged to distinct sublineages (A.1, A.2 and A.3), and the AZ1 genome contained one derived substitution (C-to-T at nucleotide position 29,031) that we did not observe in any other Arizona sequences. This substitution was, however, shared with 12 other genomes included in our analysis, all of which were sampled in China or Japan. This is consistent with infection of AZ1 occurring during documented travel to China^5^.

**Figure 1.**
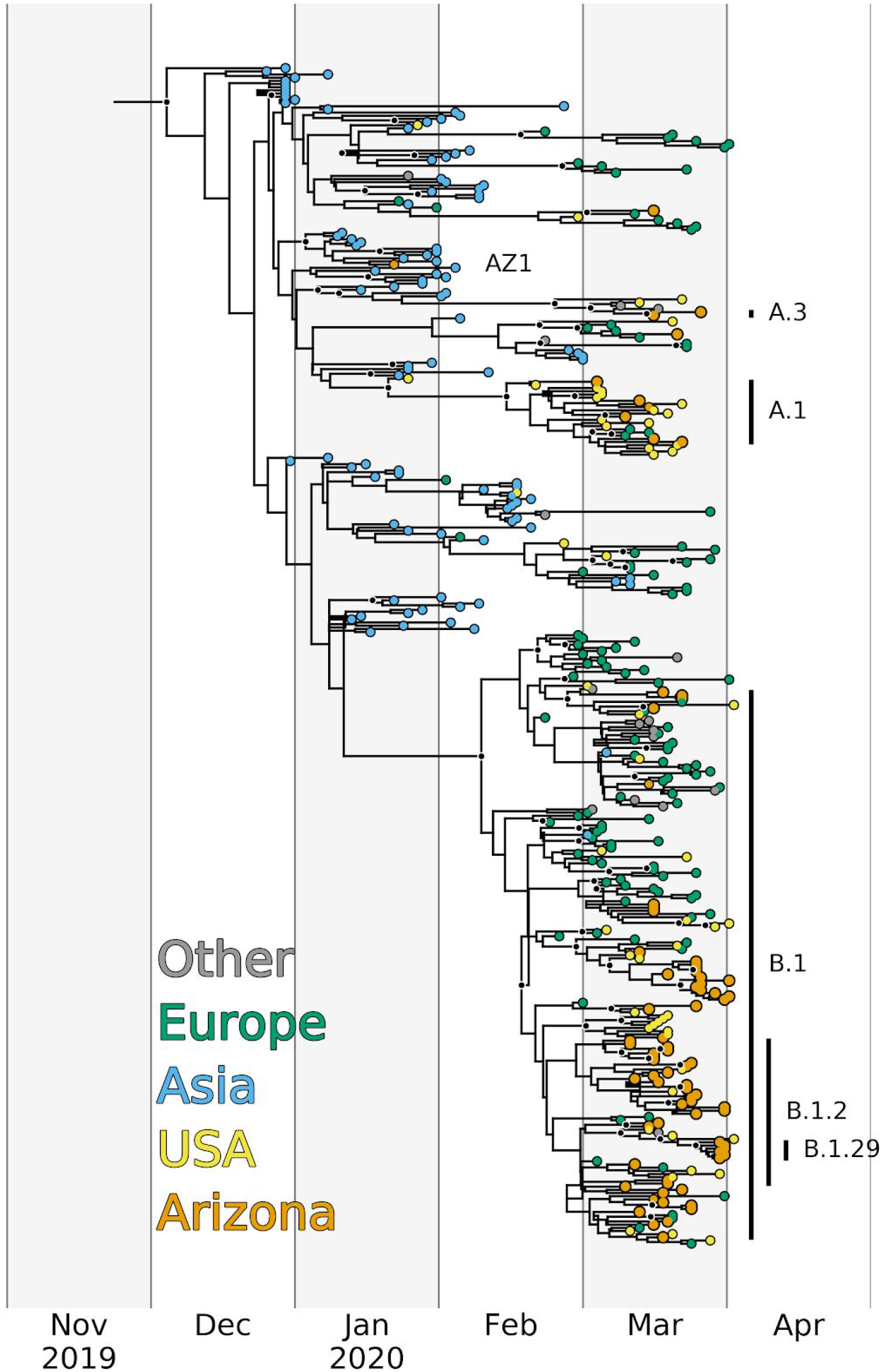
Bayesian maximum clade credibility time-calibrated phylogeny inferred from 372 SARS-CoV-2 genomes, including 80 from Arizona and 292 representatives from around the world. Tips are colored by origin of sequence, and major lineages assigned by Pangolin with more than 2 sequence representatives in Arizona are indicated by vertical bars. All nodes with posterior probabilities >0.9 are colored black. The tree was visualized with a custom Python script that utilized the software package BALTIC (https://github.com/evogytis/baltic).

Although our analyses cannot completely rule out the possibility of a cryptic transmission chain originating from this initial case, it is clear that the first documented introduction of SARS-CoV-2 to Arizona did not play a substantial role in fueling the ongoing epidemic. Rather, the majority of Arizona cases are linked to later introduction events (see below). These results further demonstrate the power of public health contact tracing and self-isolation following a positive test for stemming the tide of infections moving forward.

### Multiple introductions have contributed to transmission in Arizona

Our phylogenetic analyses indicated that multiple distinct SARS-CoV-2 lineages co-circulated in AZ during March 2020. We detected 9 distinct lineages/sublineages in Arizona between early March and early April (Table 1). These lineages were not unique to AZ; they have all been documented in other parts of the USA, as well as in multiple countries around the world (Figure 3, Supplementary Figure 2). In fact, the geographic ubiquity of the major SARS-CoV-2 lineages, along with the common observation of identical virus genomes sampled in multiple US states, countries and continents, clearly demonstrates how frequently this virus has been moved between locales. Given the timing and size of the AZ outbreak, relative to outbreaks in other locations (Figure 3), we argue it is unlikely that any of these named lineages arose within Arizona. Therefore, the number of observed lineages (10 including AZ1) represents a conservative estimate for the number of independent introductions of SARS-CoV-2 into Arizona.

**Table 1.** Information on sequence number, timing, and location of each of 10 lineages detected in Arizona.

| Lineage | Number of sequences | Date collected in AZ | Counties |
| --- | --- | --- | --- |
| A | 1 | 1/22/20 | Maricopa |
| A.1 | 6 | 3/5/2020 to 3/23/2020 | Graham, Maricopa, Mohave, Pima, Pinal |
| A.2 | 1 | 3/22/20 | Cochise |
| A.3 | 2 | 3/17/20 to 3/27/20 | Coconino |
| B.1 | 37 | 3/11/20 to 4/2/20 | Coconino, Maricopa, Navajo, Pima, Pinal, Yuma |
| B.1.2 | 24 | 3/12/20 to 4/1/20 | Coconino, La Paz, Maricopa, Navajo, Pinal, Yavapai |
| B.1.21 | 1 | 3/13/20 | Pima |
| B.1.29 | 6 | 3/13/20 to 4/1/20 | Maricopa |
| B.1.3 | 1 | 3/19/20 | Coconino |
| B.2 | 1 | 3/17/20 | Coconino |

To estimate when community transmission of SARS-CoV-2 first began in Arizona, we used our Bayesian phylogenetic analysis to estimate dates for the most recent common ancestors (tMRCA) of the Arizona genomes within each of these lineages. In total, we sequenced ≥2 viral genomes from 5/10 of the observed lineages (which we name “major lineages”), and tMRCA estimates using these genomes (Figure 2) suggest that, at the earliest, community transmission in Arizona began around early February 2020. However, we view this as the most extreme estimate because many of these lineages likely represent multiple distinct introductions to Arizona, rather than a single introduction followed by sustained community transmission. For example, B.1, the inferred oldest lineage containing Arizona genomes, includes multiple well-supported clades (posterior probability ≥ 0.99, bootstrap ≥65), all of which have also been observed in several other US states and countries (Supplementary Figure 1). Therefore, there have almost certainly been multiple introductions of viruses from this lineage that have contributed to local transmission in Arizona, and the early February tMRCA likely predates actual community transmission within Arizona..

**Figure 2.**
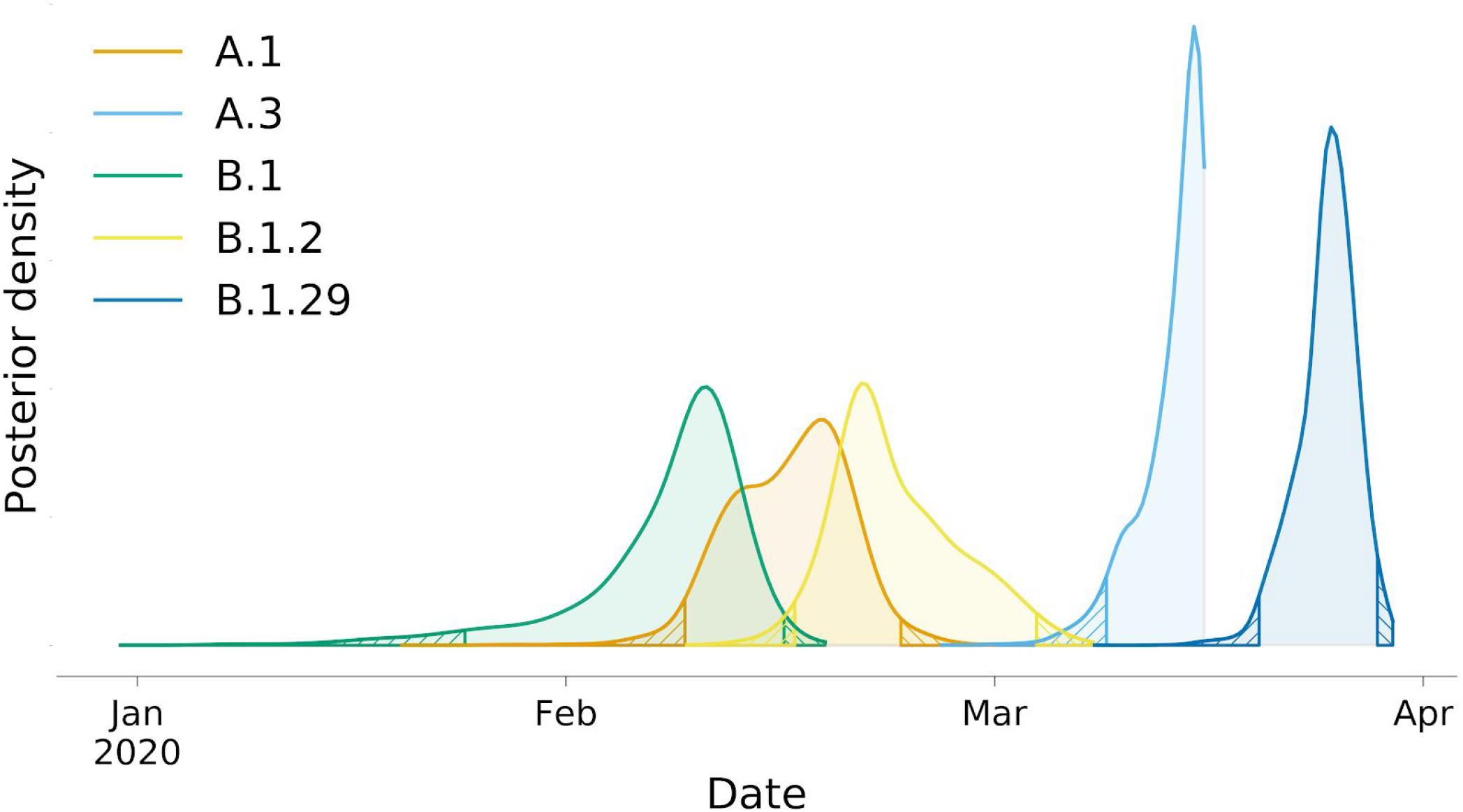
Posterior density estimates of tMRCAs for Arizona genomes that belong to 5 major lineages/sublineages. Posterior density estimates were parsed from 9001 trees sampled from 4 independent MCMC chains, following burn-in removal. Hatch marks indicate regions outside of the 95% highest posterior density (HPD).

To determine the most likely source of the SARS-CoV-2 introductions to Arizona, we examined the geographic distribution of sequences within each lineage (Figure 3). For this analysis, we considered the full collection of SARS-CoV-2 genomes available on GISAID (as of 4/16/2020). First, for each of the 5 major lineages, Arizona sequences were always sampled after a sequence from that same lineage or sublineage had been sampled elsewhere. Although sequencing efforts can influence this result, it appears unlikely that any of the major lineages emerged in Arizona; rather, they were likely imported. Second, there are very few sequences from Asia clustered within each of the 5 major lineages. Instead, it appears importation was largely driven by domestic travel (in the case of A.1, B.1.2, B.1.29), or perhaps imported from Europe (B.1) or Oceania (A.3), although both of these latter lineages have also been commonly observed elsewhere in the US. Given the widespread distribution of these nearly identical genomes, it would be impossible to directly estimate the location of each of these importations beyond the continent level.

**Figure 3.**
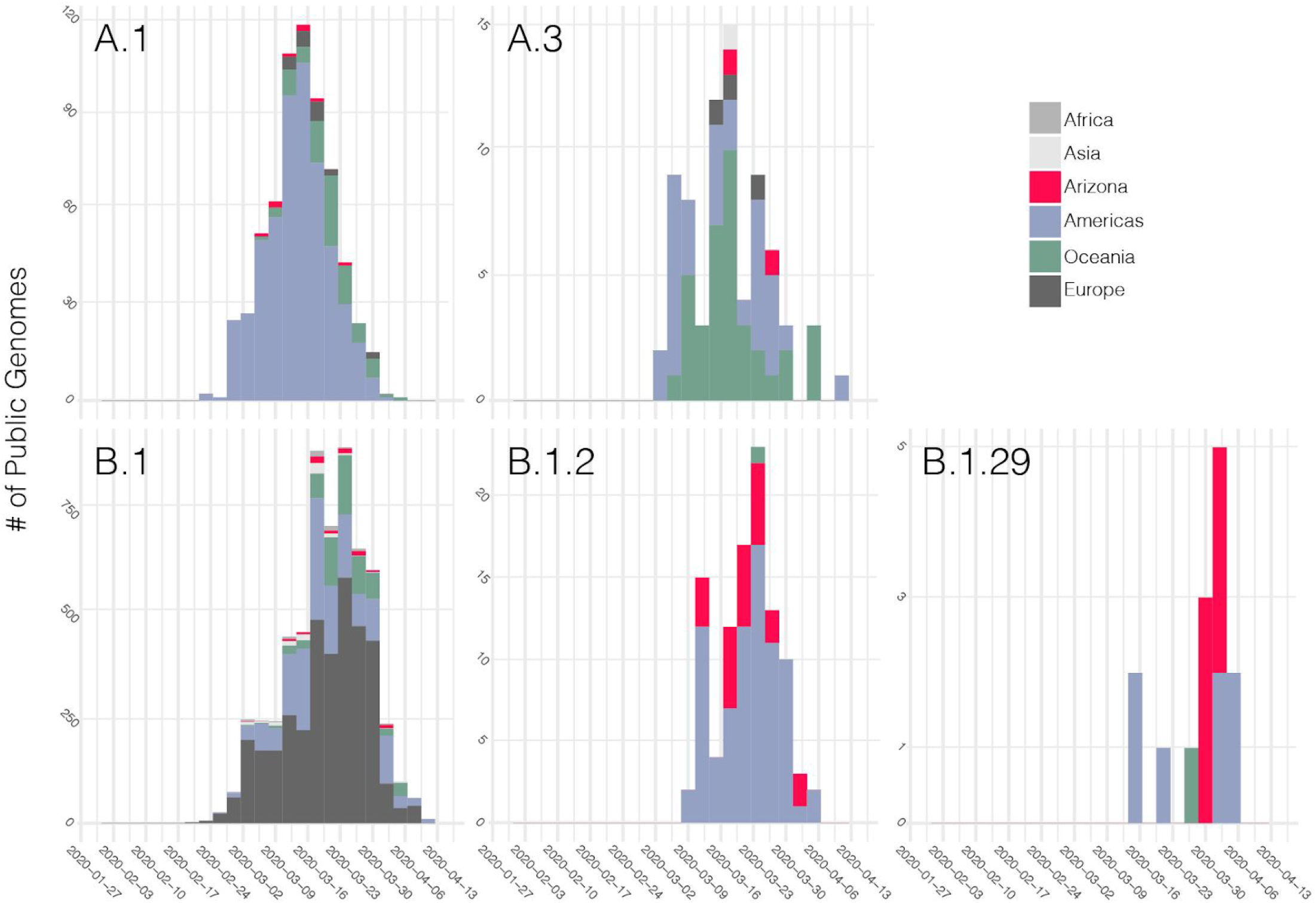
Sequence database representation through time for each of the 5 major lineages/sublineages that occur in Arizona. Stacked bars are colored according to location. To estimate A.1, B.1, and A.3, nested sublineages were collapsed in order to calculate the frequencies for the broader clades. Lineages were assigned using Pangolin for all sequences uploaded to GISAID as of 4/16/2020.

The Arizona sequences are largely represented by two lineages, A.1 and B.1. We focus the remaining results and discussion on these two lineages. A.1 was identified in Washington State in late February and has since spread across the globe (Figure 3). It was previously proposed that the A.1 Washington outbreak, announced on February 28th as the 2nd instance of community transmission in the United States^14,15^, is derived from the first Washington case, a lineage A sequence, back in January 2020^13^. For the purposes of our discussion, we only focus on the A.1 sublineage, which, regardless of the ultimate source of A.1, had begun circulating in Washington in the middle of February at the latest.

We sequenced six genomes that are members of the A.1 clade from Arizona; these were detected in 5 counties across Arizona (Table 1). We infer that the MRCA of these Arizona sequences likely existed around Feb 16th 2020 (95%HPD Feb 9th - Feb 23rd). If these six Arizona genomes stem from a single introduction of the A.1 lineage, this tMRCA estimate suggests the lineage was already present in Arizona prior to when community transmission was announced in Washington, on Feb. 28th (Figure 2). However, it is possible, and even likely, that the six A.1 lineage genomes from Arizona arose from multiple introductions into the state, given that the same lineage was being spread throughout the country^16^. Although multiple introductions would push the tMRCA estimate of the A.1 introduction to be more recent, we argue that it is possible A.1 was first imported near the dates inferred.

The first A.1 Arizona genome we sequenced was collected on March 5th from a person who was a household contact of another case, representing the first known community transmission event in Arizona; the other individual tested positive on March 3rd and held a healthcare job in Phoenix (their SARS-CoV-2 genome has not been sequenced)^17^. This implies the first community transmission subject was infected, sought testing and infected a close contact who we sampled. Given the median incubation time of 5 days,^18^ and assuming the healthcare worker was infected by someone traveling directly from Washington with no additional transmission in between, that would place the time of importation to be ~Feb. 28th. This falls outside our 95% HPD tMRCA estimate of Feb 23rd; however, this timeline makes it clear that even if multiple A.1 introductions caused the tMRCA to be artificially early, it is not by more than a week or two.

Of the 79 genomes we have sequenced in Arizona, 65 (82%) belong to the B.1 lineage (including sublineages B.1.2 and B.1.29), making B.1 the most abundant lineage in Arizona (as it is globally)^19^. Arizona sequences from this lineage were collected from March 11th to April 2nd, 2020 and were detected in samples from eight counties (Table 1). If these genomes all stemmed from a single introduction to Arizona, we estimate an introduction around Feb 7th, 2020 (95%HPD Jan 24th - Feb 15th). However, given the widespread distribution and frequency of this particular clade, both globally and nationally, as well as the lack of detected cases in Arizona in February, it is very likely that multiple introductions of B.1 viruses have contributed to the outbreak in Arizona, in which case, the actual onset of community transmission for this lineage may be later than the tMRCA estimate suggests.

One of the substitutions that is present in all B.1 lineage viruses occured in the gene for the spike protein, and resulted in an aspartic acid to glycine substitution at residue 614 (D614G). Based on viral RNA quantifications and phylogenetic analyses^20^, it was recently suggested that this substitution may have increased the transmissibility of the virus. Although the outbreak in Arizona was already dominated by B.1 lineage viruses in early March 2020, we did see a gradual increase in the relative proportion of B.1 throughout March and into early April (Figure 4A). We also compared qRT-PCR cycle threshold (Ct) values for clinical samples with and without this substitution. Our results show a similar trend to that reported from patients in Sheffield, England^20^, with a lower mean Ct (higher viral load) in samples containing the D614G substitution; however, this is not a statistically significant difference (p=0.85) with our current sample size (Figure 4B). Combined, these data, along with those published by other groups^20^ (https://github.com/blab/ncov-D614G), are consistent with a replication and/or transmission advantage of viruses containing the D614G substitution. However, experimental studies are needed to validate this hypothesis. An alternative explanation is that the B.1 lineage has simply been increasing in frequency globally, due to chance events that led this lineage to dominate in early outbreaks (e.g., in Italy), from which the virus has been spread widely throughout the world and has seeded outbreaks in many other locations.

**Figure 4.**
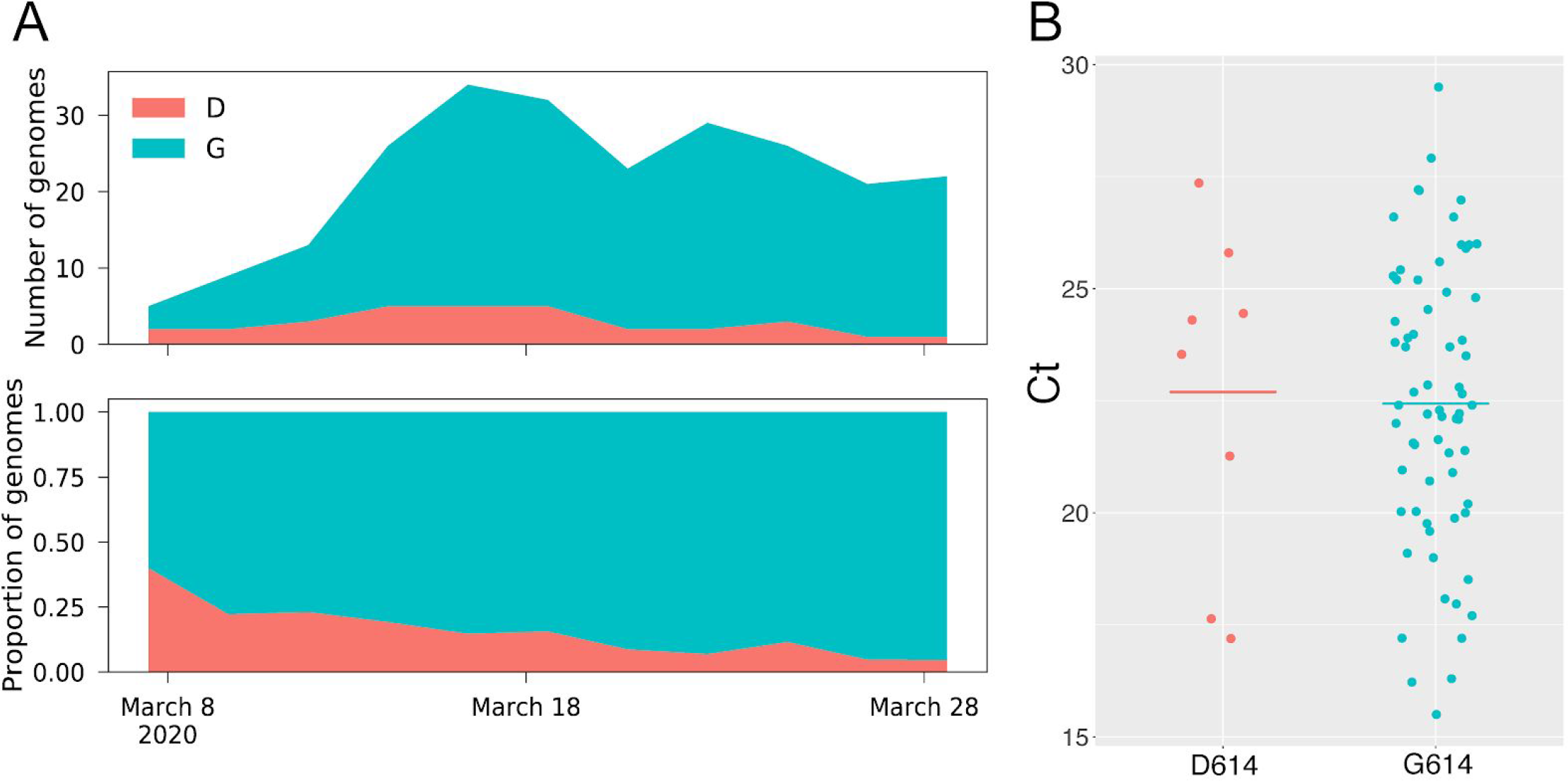
Abundance over time (A) and cycle threshold values (B) for viruses in Arizona with and without the D614G substitution. Both panels were generated using the 79 Arizona genomes we report here. Plots of abundance over time were generated using a window size of 1 week and a step size of 2 days.

The B.1 lineage has also dominated the large outbreak in New York, and in some reports^21^, the B.1 lineage has been used as an indication of an epidemiological link to the New York outbreak. However, we would urge caution with this approach. Based on ancestral state reconstructions, this lineage is predicted to have first emerged in Asia and/or Europe (Nextstrain), and this lineage was observed in multiple European countries, including the large Italian outbreak, before it was first documented in New York (Nextstrain). In fact, just within Arizona, we have documented at least two instances in which B.1 lineage viruses were imported directly to the US from Europe. The second case of COVID-19 in Arizona was reported on March 3rd, from a traveler who returned from France on Feb. 27th^22^. This individual self-reported being symptomatic on the plane back to Phoenix, and went to multiple social gatherings before being officially diagnosed^23^. At one of these social events, at least one other individual was infected^24^. Although we did not sequence the SARS-CoV-2 genome from this individual, CDC deposited this sequence into GISAID (EPI_ISL_420784), and we confirmed it belonged to the B.1 lineage (data not shown). We also have records shared to us by the Coconino Health and Human Service Department that one of the B.1 genomes we sequenced from Coconino County came from an individual who had recently traveled to Rome, Italy, and who presumably got infected there. Thus, although it is tempting to speculate that most of the B.1 infections across the US came from the NY outbreak, we show at least two confirmed instances of direct importation of B.1 into Arizona from Europe.

### Non-synonymous mutations of interest observed in Arizona SARS-CoV-2 sequences

Like all RNA viruses, SARS-CoV-2 accumulates mutations over time, some of which may impact virulence, replication and intervention strategies, and some of which have no functional, clinical or antigenic importance. We identified non-synonymous mutations in coding sequences of SARS-CoV-2 genomes from Arizona (Figure 5, Supplementary Table 2). These include mutations in the spike protein, nonstructural proteins (nsp) involved in RNA synthesis, nucleocapsid protein, and the putative ORF10. Several of these mutations have also been reported as being associated with SARS-CoV-2 sequences from Europe^25^. Below, we hypothesize about potential phenotypic impacts of these substitutions; however, it is important to note that experimental studies need to be conducted to test these hypotheses, and the vast majority of substitutions that occur during viral replication will not have a significant impact on virulence or transmissibility.

**Figure 5.**
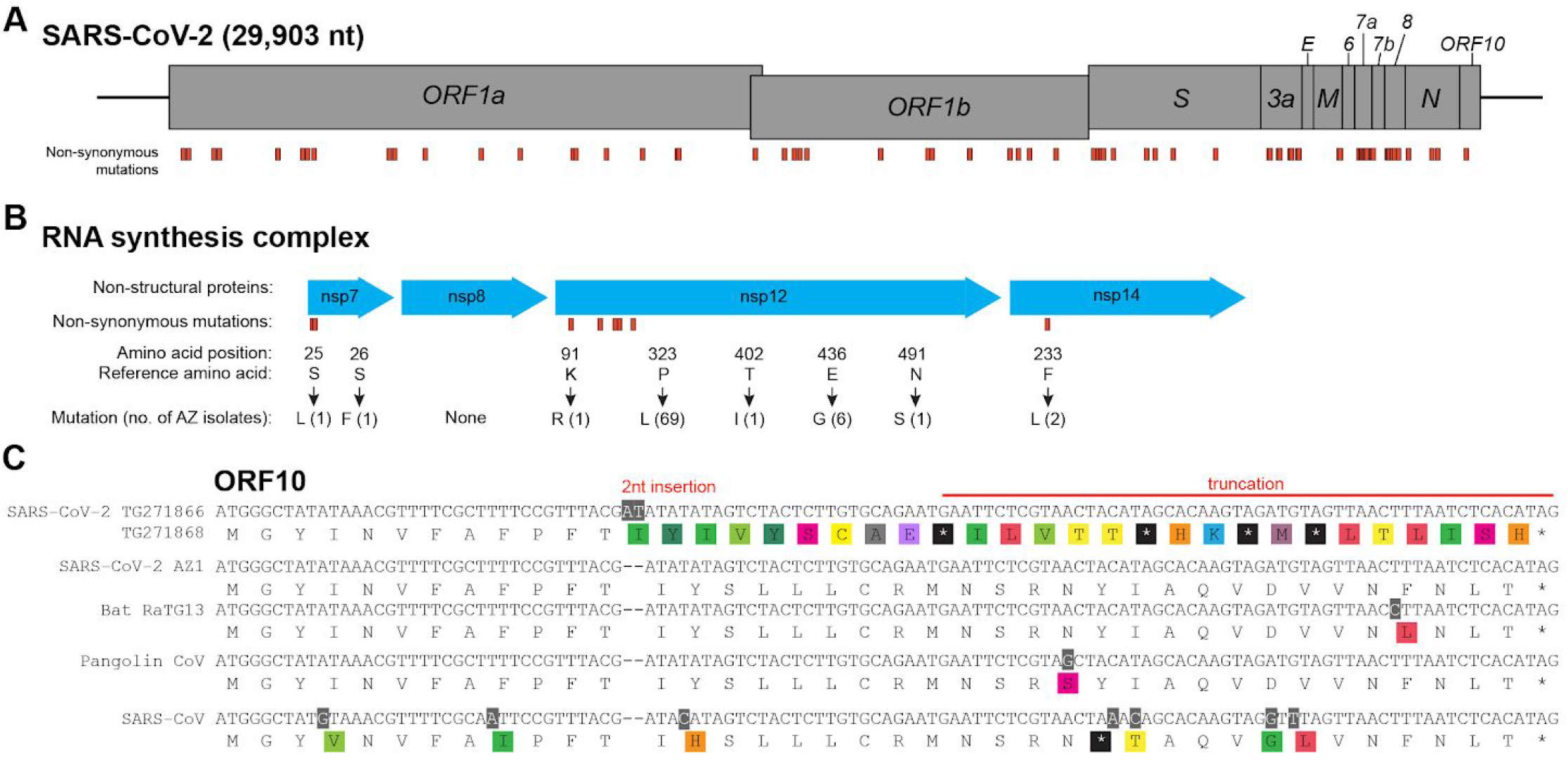
Non-synonymous mutations in Arizona isolates. (A) Diagram shows SARS-CoV-2 genome and annotated open reading frames. Genome position of non-synonymous mutations in Arizona SARS-CoV-2 isolates are indicated in orange. (B) Non-synonymous mutations of Arizona isolates in nsp involved in the SARS-CoV-2 RNA synthesis complex. Mutations (indicated in orange) are labeled by amino acid position within the protein, reference amino acid, amino acid change, and number of Arizona isolates with the mutation. (C) ORF10 alignment showing 2-nucleotide insertion and subsequent early truncation in two Arizona SARS-CoV-2 isolates. GenBank and GISAID accession numbers: SARS-CoV-2 AZ-TG271866 (EPI_ISL_427271), SARS-CoV-2 AZ-TG271868 (EPI_ISL_427272), SARS-CoV-2 AZ1 (MN997409.1, EPI_ISL_406223), Bat-RaTG13 (MN996532.1), Pangolin (EPI_ISL_410721), SARS-CoV (NC_004718.3).

To identify if any mutations could affect the specificity of currently used qRT-PCR assays, an *in silico* approach was employed. A screen of 12 current primer/probe sets (Supplementary Table 4) demonstrates that 7 assays yield no false negatives based on exact primer/probe matches. Several of the primers/probes align with stretches of “N” characters in individual genomes and the amplification potential is therefore ambiguous. Interestingly, the CDC nCoV_N2 assay demonstrates mismatches in 6 of the genomes screened in this study, including one genome from Arizona (TG271862, Cochise County).

#### Spike

The SARS-CoV-2 spike (S) protein mediates receptor binding and cell entry, and is the primary target of neutralizing antibodies^26^. Mutations in the spike protein may have implications for viral entry and recognition by the immune system. We found several mutations in the spike protein gene, including two epidemiologically linked isolates (TGEN-CoV-AZ-WMTS-TG268282 and TGEN-CoV-AZ-WMTS-TG271435) harboring an alanine to valine substitution at Spike amino acid residue 475 (A475V, nt position 22986) in the receptor binding domain (RBD). Structural studies indicate that A475 interacts with S19 of ACE2^27,28^.

#### Nsp12 and RNA synthesis complex

During replication of the SARS-CoV-2 RNA genome, RNA synthesis is driven by the key component nsp12 RNA-dependent RNA polymerase (RdRp) in complex with nsp7 and nsp8^29,30^. Studies of SARS-CoV replication demonstrate that nsp14 regulates replication fidelity through its 3’-to-5’ exonuclease activity^31,32^. Mutations in nsp7, nsp8, nsp12, and nsp14 may therefore have an effect on viral RNA synthesis and susceptibility to antiviral treatments such as remdesivir^33^. We identified several non-synonymous mutations in nsp7 residue 25 (S → L) and 26 (S → F), but none in nsp8 (Figure 5B). One of the nsp12 mutations at residue 323 (P → L) identified in 69 AZ sequences was previously associated with SARS-CoV-2 sequences from Europe^25^. We did not find nsp12 mutations at sites predicted to be the contact interface with remdesivir^33^. Finally, the 2 isolates with nsp14 mutation at residue 233 (F → L) were the same sequences harboring the spike RBD A475V mutation.

#### Nucleocapsid

The nucleocapsid (N) protein encapsidates the genomic RNA and is a target for diagnostic and therapeutic applications^34^. The N protein is also associated with replication-transcription complexes and facilitates template switching during viral subgenomic mRNA synthesis^35^. N is expressed at high levels during early stages of replication and, like the S protein, is also a major immunogenic target of antibodies^36^. Five AZ sequences had a triple nucleotide substitution (GGG → AAC) that resulted in a tandem amino acid change in the N protein at residues 203 and 204 (RG → KR). Over the relatively short time-frame of SARS-CoV-2 evolution, these tandem substitutions are relatively uncommon^25^.

#### ORF10

ORF10 is a short putative protein of unknown function, predicted in the 3’ end of the SARS-CoV-2 genome, which is conserved in the closely-related bat and pangolin coronavirus sequences (Figure 5C). We identified a 2-nucleotide insertion in ORF10 from two AZ sequences that result in a premature stop codon and early truncation of ORF10. A similar truncation is present in the SARS-CoV genome due to the presence of an upstream stop codon. This may indicate a region of the virus genome with relaxed evolutionary constraints, consistent with a report that did not detect subgenomic ORF10 mRNA^37^.

## Conclusions

Based on our phylogenetic analysis, it is clear that the ongoing COVID-19 outbreak in Arizona has been fueled by multiple distinct introductions of SARS-CoV-2 to the state. We estimate a minimum of 9 over the course of February and March. By estimating the timing of introductions, we find no evidence for cryptic community transmission in Arizona prior to late January. Rather, our analyses indicate that community transmission likely did not occur within Arizona until at least early-to mid-February when viruses from lineages B.1 and A.1 may have been introduced. It appears that most of the introductions of SARS-CoV-2 to Arizona have had domestic origins, in line with reports from other states^16^; however, there have also been instances of Arizona cases linked to international travel, and these have likely also contributed to the local outbreak. A number of non-synonymous mutations were identified in the Arizona isolates, including within regions of the receptor-binding domain of the spike protein and non-structural proteins involved in the RNA synthesis complex. The functional consequences of these mutations are unknown, highlighting the need for mechanistic studies. Our phylodynamic tracing provides unique epidemiological insights into the origins and transmission of SARS-CoV-2 in Arizona.

## Methods

### Reference genome

Any sequence positions mentioned in this work refer to GenBank sequence NC_045512.2, a genome isolated and sequenced from Wuhan early in the pandemic.

### TGen North Genome Sequencing

RNA was extracted from specimen transport medium with the Quick Viral RNA kit (Zymo Research). Total RNA sequencing libraries were prepared with the SMART-Seq® Stranded Kit (Takara) or the Ovation RNA-Seq System (NuGEN). Libraries were sequenced on a NextSeq (high output kit, Illumina). Viral genome consensus sequences for each sample were constructed using TGen’s Amplicon Sequencing Analysis Pipeline (ASAP, https://github.com/TGenNorth/ASAP) which consists of mapping the reads to a reference genome (hCoV-19/USA/AZ1/2020|EPI_ISL_406223^38^) and analyzing the alignment pileup position by position to determine coverage depth, coverage breadth and the consensus sequence. Positions covered by fewer than 10 reads were considered a gap in coverage and were converted to Ns, Consensus sequences were saved and used for further analysis when they had >90% breadth of coverage and >30x average depth of coverage.

### University of Arizona Genome Sequencing

Following sample collection, the nasopharyngeal swab was soaked in TRIzol (ThermoFisher) and removed. Total RNA was then extracted from 400uL of TRIzol with the Direct-zol RNA isolation kit (Zymo Research) following the manufacturer’s instructions. RNA was eluted in 30uL of nuclease-free water.

We used primers and methods from the ARTIC consortium^39^ with the following modifications. cDNA synthesis was performed with GOscript (Promega) using 10uL of RNA in a final volume of 20uL following manufacturer’s instructions. Next, a multiplex PCR amplifying overlapping 400bp amplicons was done with the V2 set of primers designed by the ARTIC group. We used 2.5uL of cDNA in each 25uL reaction with the Q5 Hot Start High-Fidelity DNA Polymerase (NEB), with 2 separate reactions containing each of the non-overlapping primer pools. We used an initial denaturation step of 98°C for 30 seconds and then 35 cycles at 98°C for 10 seconds, 65°C for 2 minutes.

The reactions were pooled together, and visualized on a 2% agarose gel to confirm successful amplification. Amplicons were then cleaned with a 1:1 mixture of AMPure XP magnetic beads (Beckman Coulter). The mixture was incubated for 5 minutes at room temperature and then placed on a magnetic rack until all the beads were pulled out of solution. The remaining liquid was pulled off and discarded. Next, 200uL of 80% EtOH was used twice to wash the bead mixture. The beads were allowed to dry and re-suspended in 30uL of water. Following a 5 minute incubation the tube was placed back on a magnetic rack until the beads were pulled out of the solution. The 30uL of cleaned amplicons was transferred to a fresh tube.

The remainder of the protocol was identical to the ARTIC protocol which includes end-repair, ligation of Oxford Nanopore sequencing adapters, and additional cleaning steps using the AMPure XP beads. The final prepared library was loaded onto a flongle inserted into a MinION sequencer. Sequence data were collected for 12 hours. In order to remove the sequencing adapter and primer sequence we trimmed the first 40bp off of the reads. Reads were then aligned to a SARS-CoV-2 reference sequence (MN908947) using Geneious Prime (Biomatters Inc.). A consensus sequence was generated from these reads for sections that contained >40x coverage with Ns placed at sites with lower coverage.

### Arizona State University Genome Sequencing

SARS-CoV-2 genomes were sequenced from nasopharyngeal swabs as previously described^40^. Briefly, total nucleic acid was extracted using the bioMérieux eMAG platform. RNA was subjected to Ribo-Zero Gold depletion, TruSeq RNA library preparation, and sequenced on Illumina NextSeq (2×76). Sequencing reads were quality filtered with BBtools (BBMap – Bushnell B. – sourceforge.net/projects/bbmap/) and mapped to a SARS-CoV-2 reference genome (MN908947).

### Bioinformatics

Sequences included in the analyses presented here were derived from GISAID (accessed on April 16th, 2020), NCBI GenBank, and sequences generated by our teams at Northern Arizona University and TGen North (n=75), Arizona State University (n=3), and University of Arizona (n=1) (collectively referred to as the “Arizona sequences”). To support efficient Bayesian phylogenetic analysis of this large number of sequences, we developed a novel protocol and software for sampling sequences from GISAID across time of sequence acquisition, geographic source of sequence, and SARS-CoV-2 diversity. The software developed for this workflow is available at https://github.com/caporaso-lab/az-covid-1, and our application of this workflow is detailed in our Supplementary Methods.

This protocol works as follows:

1. Download SARS-CoV-2 sequences from GISAID (9230 at time of download)
2. Clean-up and filter GISAID sequences (8659 sequences remained after these filters)

a. Specific steps were:

i. Filter sequences that contain spaces (spaces are non-IUPAC characters^41^) - this resulted in 408 sequences being discarded.
ii. Remove any gap (“-”) characters (sequences are unaligned, so should not contain gaps)
iii. Filter sequences that are composed of >10% N characters - this resulted in 163 sequences being discarded.
iv. Replace all spaces in sequence header lines with underscores
3. Compile initial sequence collection containing Arizona sequences, reference sequence from NCBI GenBank (NC_045512.2),seven randomly sampled sequences per week from GISAID, WA1 (MN985325.1), and AZ1 (MN997409).
4. We then searched all of GISAID against the sequence collection compiled in Step 3 using vsearch’s usearch_global option, and identified sequences that most closely matched the Arizona sequences, to ensure that we were including the closest relatives of Arizona sequences to contextualize clades containing Arizona sequences ^42^. The collection of best hits was sampled to result in a geographically dispersed collection of sequence sources. Those sequences were added to the sequence collection from Step 3, and ensure that monophylies of Arizona sequences are not artifacts of our sequence sampling approach.
5. All GISAID sequences that were not clustered by the previous step were clustered with vsearch’s cluster_fast option at 99.9% percent identity. Cluster centroid sequences were added to the sequence collection generated in step 4. This ensured that a divergent collection of the SARS-CoV-2 genomes was represented in our data set.
6. An initial alignment was constructed with MAFFT followed by the construction of a neighbor joining tree. Several sequences that were suspected of being low quality at this stage, including both GISAID and Arizona sequences, were removed from the sequence collection.

The sequences resulting from this workflow were used as the starting point for all downstream analyses. Maximum-likelihood phylogenies were generated using RAxML-NG v0.5.1b^43^ with the GTR+G4 model, as indicated by a substitution model selection analysis carried out in IQTree, with 20 distinct starting trees and 100 bootstrap replicates. Lineage assignment for all Arizona sequences was performed with the software Pangolin, which is a real time dynamic tool for assigning lineages to SARS-CoV-2 sequences based on shared mutations and phylogenetic support^44^.

We employed a Bayesian molecular clock method implemented in the BEAST v1.10.4^45^ software package to estimate the divergence times for the total SARS-CoV-2 dataset as well as several Arizona-specific lineages, and overall evolutionary rates. To compare how different model combinations impact results, we carried out two sets of analyses with the Uncorrelated Lognormal (UCLN)^46^ clock model combined with either the Exponential or Bayesian Skygrid demographic^47^ models. Four chains of each model combination were run for 100,000,000 generations and sampled every 10,000 generations. For both model combinations, we found convergence within and among chains using Tracer v1.6. LogCombiner was used to merge the four different chains of each model combination, after discarding the first 10% as burn in (10,000,000 generations per chain), and then resampling every 40,000 generations. The two resulting files were input to TreeAnnotator to produce maximum clade credibility trees with median height estimates. BALTIC (https://github.com/evogytis/baltic) was used for visualizing trees and for parsing tMRCAs from the trees sampled during the BEAST analysis.

The in silico PCR screen was performed with an *in silico* PCR script (https://github.com/TGenNorth/vipr) using published primers and probes (Supplementary Table 4). Positives were identified by exact nucleotide matches of both primers and probe. Mishits were manually identified through visual inspection of the primer/probe alignments. If a primer/probe aligned against a genome with an “N” character, the hit was considered as ambiguous. The Arizona SARS-CoV-2 isolates were aligned against reference genome AZ1 (GenBank accession MN997409, GISAID accession EPI_ISL_406223) and non-synonymous variants in the coding sequences were called by Geneious Prime (2020.0.5).

## Data Availability

All sequences have been deposited into the GISAID repository. The software developed for our sequence collection sampling workflow is available at https://github.com/caporaso-lab/az-covid-1 under the BSD 3-clause license. Supplementary Tables 1 and 2 can be found at https://github.com/caporaso-lab/az-covid-1/tree/preprint.v1/paper1

https://github.com/caporaso-lab/az-covid-1

https://github.com/caporaso-lab/az-covid-1/tree/preprint.v1/paper1

## Acknowledgements

We are deeply indebted to all the patients who provided samples (via an uncomfortable procedure) while suffering from this new pathogen, and to all the medical professionals who bravely collected samples that form the basis for this work. Furthermore, we would like to acknowledge the GISAID sequence contributors from across the world who have sequenced genomes at an unprecedented rate and submitted them for public use. We would especially like to thank the CDC group that sequenced the first Arizona COVID-19 case (Ying Tao, Clinton R. Paden, Krista Queen, Anna Uehara, Yan Li, Jing Zhang, Xiaoyan Lu, Brian Lynch, Senthil Kumar K. Sakthivel, Brett L. Whitaker, Shifaq Kamili, Lijuan Wang, Janna’ R. Murray, Susan I. Gerber, Stephen Lindstrom, Suxiang Tong). This work was supported in part by funds provided by The NARBHA Institute, The Flinn Foundation, The Virginia G. Piper Charitable Trust, and Blue Cross and Blue Shield of Arizona (DME, JRB) as well as the National Institutes of Health (NIH grant: R00 DK107923) (ESL), and David and Lucile Packard Foundation as well as the University of Arizona College of Science, BIO5 Institute and Office of Research Innovation and Impact (MW). We would also like to acknowledge the critical role the Arizona Department of Health Services and multiple county health departments played in directing samples to us to be sequenced. Computational analyses were run on Northern Arizona University’s Monsoon computing cluster, funded by Arizona’s Technology and Research Initiative Fund. Additional analysis effort was funded under the State of Arizona Technology and Research Initiative Fund (TRIF), administered by the Arizona Board of Regents, through Northern Arizona University. The Cowden Endowment for Microbiology provided funds to support salaries.

## Author Contributions

Conceptualization: JTL, BBL, JRB, PK, MW, DME, ESL

Data curation: BBL, JRB, JWS, DL, AP, HY

Formal Analysis: JGC, BBL, JTL, CMH, RM, DL

Funding acquisition: PK, DME, MW

Investigation: BBL, GQ, TDW, JTL, CMH, EAK, ESL, JWS, JRB, MF, DME, MW

Methodology: JTL, CMH, MF, KS, AP, DL

Project administration: PK, MW, DME, JRB

Resources: KK, VW, DME

Software: JGC, EB, NAB

Supervision: PK, DME, MW

Validation: MF, KS, DL

Visualization: BBL, JTL, EAK, RM

Writing - original draft: JGC, BBL, JTL, CMH, EAK, ESL, JRB, HY

Writing - review & editing: JGC, BBL, JTL, CMH, PK, JWS, ESL, JRB, DME, NAB, MW

